# Cardiovascular Risk Factors and Outcomes in COVID-19: Hospital-Based Prospective Study in India

**DOI:** 10.1101/2021.09.19.21263788

**Authors:** Arvind Sharma, Vaseem Naheed Baig, Sonali Sharma, Gaurav Dalela, Raja Babu Panwar, Vishwa Mohan Katoch, Rajeev Gupta

## Abstract

**Background & Objectives:** Presence of cardiovascular (CV) risk factors enhance adverse outcomes in COVID-19. To determine association of risk factors with clinical outcomes in India we performed a study.

**Methods:** Successive virologically confirmed patients of COVID-19 at a government hospital were recruited at admission and in-hospital outcome and other details obtained. The cohort was classified according to age, sex, hypertension, diabetes and tobacco use. To compare intergroup outcomes we performed univariate and multivariate logistic regression.

**Results:** From March-September 2020 we recruited 4645 (men 3386, women 1259) out of 5103 COVID-19 patients (91.0%). Mean age was 46±18y, hypertension was in 17.8%, diabetes in 16.6% and tobacco-use in 29.5%. Duration of hospital stay was 6.8±3.7 days, supplemental oxygen was in 18.4%, non-invasive ventilation in 7.1%, mechanical ventilation in 3.6% and 7.3% died. Unadjusted and age-sex adjusted odds ratio and 95% confidence intervals, respectively were, age ≥50y (4.16, 3.22-5.37 and 4.15,3.21-5.35), men (1.88,1.41-2.51 and 1.26,0.91-1.48); hypertension (2.22,1.74-2.83 and 1.32,1.02-1.70), diabetes (1.88,1.46-2.43 and 1.16,0.89-1.52) and tobacco (1.29,1.02-1.63 and 1.28,1.00-1.63). Need for invasive ventilation was greater in age >50y (3.06,2.18-4.28 and 3.06,2.18-4.29) and diabetes (1.64,1.14-2.35 and 1.12,0.77-1.62). Non-invasive ventilation was more in age ≥50y (2.27,1.80-2.86 and 2.26,1.79-2.85) and hypertension (1.82,1.41-2.35 and 1.29,0.99-1.69). Multivariate adjustment for presenting factors attenuated the significance.

**Conclusion:** Cardiovascular risk factors-age, male sex, hypertension, diabetes and tobacco-are associated with greater risk of death and adverse outcomes in COVID-19 patients in India.

## INTRODUCTION

Presence of cardiovascular risk factors lead to adverse outcomes in COVID-19. Studies have highlighted importance of risk factors-obesity, diabetes, hypertension, smoking and sedentary lifestyle and presence of clinical cardiovascular disease.^1^ Presence of these factors leads to rapid progression of clinical manifestations, more severe pulmonary disease, greater requirement for oxygen and ventilatory support and greater mortality.^1,2^ It has been reported that presence of hypertension, diabetes and cardiovascular disease is associated with a two-fold increase in risk of severe complications and death in COVID-19.^2^ In a meta-analysis of 109 studies and 20,296 patients, the risk of mortality was higher in patients with increasing age, male sex (relative risk, RR 1.45, 95% confidence intervals, CI, 1.23-1.71), diabetes (1.59, 1.41-1.78), hypertension, tobacco use and congestive heart failure (4.76, 1.34-16.97).^2^ In another meta-analysis of 45 studies with 18,300 patients a significant association of in-hospital death was observed with age (coefficient 1.06, 95% CI 1.04-1.09), diabetes (1.04, 1.02-1.07) and hypertension (1.01, 1.01-1.03), but remained significant only for diabetes after statistical adjustment.^3^ Another meta-analysis of 51 studies and 48,317 patients, mostly from high and upper middle income countries, reported relative risk of developing severe disease or deaths as significantly higher in patients with hypertension (RR 2.50, 95% CI 2.15-2.90), diabetes (2.25, 1.89-2.69) and cardiovascular disease (3.11, 2.55-3.79); the risk being significantly greater in older than younger individuals.^4^ Population-based studies have identified importance of cardiovascular risk factors and disease in COVID-19 incidence and outcomes.^6,7^

Burden of COVID-19 related mortality is high in lower-middle and low-income countries of South Asia and Africa.^8^ Prevalence of cardiovascular risk factors is high in these countries.^9^ India has high burden of cardiovascular risk factors,^9^ and COVID-19 cases and deaths.^10^ There are limited data on association of cardiovascular risk factors with disease incidence and outcomes in India.^11-13^ A macrolevel study in India reported that states with higher prevalence of cardiovascular risk factors-aging, hypertension, diabetes and obesity-had significantly higher COVID-19 incidence and deaths.^14^ Although higher age is well-known COVID-19 risk factor, controversy exists regarding relative importance of risk factors-hypertension, diabetes or tobacco use.^2^ Therefore, to evaluate association of cardiovascular risk factors (age, male-sex, hypertension, diabetes and tobacco use) in virologically confirmed COVID-19 cases successively admitted to a government hospital in India we performed a prospective registry-based study.

## METHODS

We conducted a hospital based prospective observational study on patients with laboratory confirmed COVID-19 admitted to a 1200-bed dedicated COVID-19 government hospital from April to mid-September 2020. Initial data on some of these patients have been reported earlier.^15-17^ The registry has been approved by the college administration and institutional ethics committee (CDSCO Registration Number CR/762/Inst/RJ/2015). It is registered with Clinical Trials Registry of India at www.ctri.nic.in with number REF/2020/06/034036.

### Patient data

Successive patients presenting to the hospital for admission with suspicion of COVID-19 infection were enrolled in the study and those who tested positive for COVID-19 on nasopharyngeal and oropharyngeal reverse transcriptase-polymerase chain reaction (RT-PCR) test were included. Details of methodology have been reported.^16^ A questionnaire was developed and details of sociodemographic, clinical, laboratory, treatments and outcomes variables were recorded using patient-history and medical files. Current smokers and users of smokeless tobacco were categorized as tobacco use. Hypertension and diabetes were diagnosed from history of known disease or on treatment. We could not obtain details of body mass index as height and weight was not routinely recorded on admission.

### Statistical analyses

The data were computerized and processing was performed using commercially available statistical software, SPSS v.20.0. Numerical data are expressed as numbers ±1 SD and categorical data as percent. Significance of intergroup differences were calculated using unpaired t-test for continuous variables and χ^2^ test for categorical variables. To evaluate association of COVID-19 related adverse outcomes (death, invasive ventilation, non-invasive ventilation) with age, male sex, hypertension, diabetes and tobacco use, we performed a stepwise logistic regression analysis. Univariate and multivariate odds ratios (OR) and 95% confidence intervals (95% CI) were calculated. In the first step we calculated univariate odds ratio. Age- and sex-adjusted odds ratios were calculated in the second step. For multivariate adjusted odds ratios we added household size, educational status, comorbidities, risk factors (other than the risk factor in question) and clinical severity (oxygen level at admission, need for oxygenation) and calculated OR and 95% CI. P value of <0.05 is considered significant.

## RESULTS

Data were obtained from March to mid-September 2020.^16,17^ During this period, a total of 7349 patients were hospitalized with confirmed or suspected COVID-19, 5103 patients (69.0%) tested positive for the disease on RT-PCR test and for the present study 4645 individuals (91.0% of confirmed cases), men 3386 (72.9%) and women 1259 (27.1%), in whom detailed clinical data were available have been included. The mean age of the cohort was 46±18 years, 54% were less than 50 years and about half lived in large family households. Prevalence of low educational status was higher in women while tobacco use was more in men. Comorbidities were present in 28.6% with hypertension (17.8%) and diabetes (16.6%) being the most common. Other comorbidities were chronic obstructive pulmonary disease (COPD), tuberculosis, coronary heart disease and neurological disease (Figure 1). Data on hematological investigations were available in 4456 (95.9%) and for biochemical tests in 867 (18.7%). All patients received standard treatment according to guidelines available from Indian Council of Medical Research and the State government.^18^ The average length of stay in hospital was 6.8±3.7 days. Oxygen requirement was in 861 (18.4%), non-invasive ventilation or high flow oxygen in 334 (7.1%) and mechanical ventilation in 169 (3.6%). In hospital mortality was in 340 patients (7.3%).

**Figure 1:**
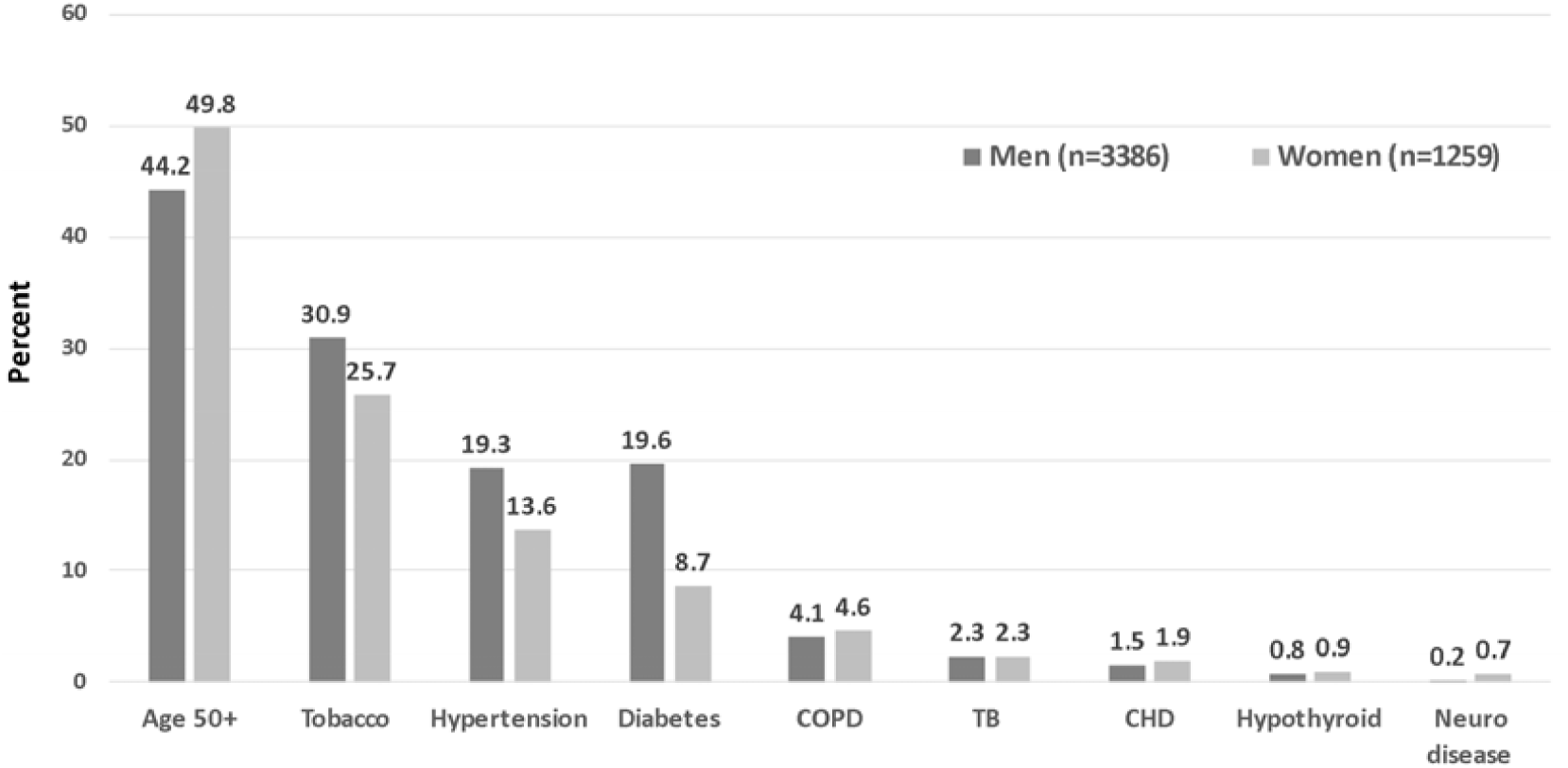
Distribution of cardiovascular risk factors among men and women in the study cohort.

Clinical characteristics, important clinical findings, selected investigations and outcomes in patients aged ≥50 years and < 50 years and in men and women are shown in Tables 1 and 2. Among patients aged ≥50y, illiteracy and low educational status, tobacco use, hypertension, diabetes, and cardiovascular disease were more and admission BP significantly higher (Table 1). In this group, compared to age <50, the need for non-invasive ventilation (OR 2.27, CI 1.80-2.86), invasive ventilation (OR 3.06, CI 2.18-4.28) as well as deaths (OR 4.16, CI 3.22-5.37) was higher. Table 2 shows that women were less literate and had lower prevalence of hypertension, diabetes and tobacco use. Oxygen requirement was significantly more in women but other outcomes such as requirement of high flow oxygen, non-invasive or invasive ventilation were not different. Number of in-hospital deaths were significantly more in men (n=282, 8.3%) as compared to women (n=58, 4.6%) (OR 1.88, 95% CI 1.41-3.51, p<0.001).

**Table 1:** Clinical characteristics and outcomes in older (>=50y) and younger (<50y) age-groups.

| | Age $\geq 50$ (n=2125) | Age- $< 50$ (n=2551) | Odds Ratio & Mean Difference (95% CI) in age $\geq 50$/$< 50$y |
| --- | --- | --- | --- |
| Age: mean±SD | 62.22±0.08 | 31.98±10.76 | -30.24[-30.81—29.66]*** |
| Low educational status | 825[38.8] | 599[23.9] | 2.07[1.82-2.35]*** |
| Household: mean±SD | 4.77±2.75 | 4.97±2.86 | 0.19[0.4-0.36]* |
| **Risk factors** |  |  |  |
| Smoking/tobacco | 648[30.5] | 721[28.3] | 1.11[0.98-1.26] |
| Hypertension | 658[31.0] | 173[6.8] | 6.16[5.15-7.38]*** |
| Type 2 Diabetes | 594[28.0] | 183[7.2] | 5.02[4.20-5.99]*** |
| Thyroid disease | 28[1.3] | 10[0.4] | 3.39[1.64-7.00]*** |
| Coronary heart disease | 60[2.8] | 15[0.6] | 4.91[2.71-8.68]*** |
| Pulmonary disease | 50[2.4] | 143[5.6] | 0.41[0.29-0.56]*** |
| **Clinical findings** |  |  |  |
| Systolic BP mmHg | 128.7±13.05 | 122.2±9.28 | -6.52[-7.16—5.88]*** |
| SpO2 $< 90\%$ | 236[11.5] | 250[10.1] | 1.15[0.95-1.39] |
| SpO2 90-94% | 332[16.2] | 369[14.8] | 1.09[0.93-1.29] |
| **Investigations** |  |  |  |
| Haemoglobin, g/dl | 12.6±2.3 | 12.8±2.3 | 0.14[-0.12-0.37] |
| White cells, $10^9$ cells/L | 7.52±3.96 | 7.53±3.72 | 0.01[-0.21-0.23] |
| Lymphocyte:neutrophil | 0.36±0.32 | 0.36±0.33 | 0.00[-0.02-0.02] |
| Serum AST, units | 40.4±35.06 | 48.0±121.86 | 7.61[-5.47-20.69] |
| Sodium, mEq/L | 136.9±6.72 | 135.3±15.82 | -1.65[-3.59-0.29] |
| Creatinine, mg/dl | 0.94±0.42 | 0.95±0.55 | 0.01[-0.06-0.08] |
| **Clinical Outcomes** |  |  |  |
| Oxygen requirement | 409[19.2] | 452[17.7] | 1.12[0.95-1.28] |
| Non-invasive ventilation | 214[10.1] | 120[4.7] | 2.27[1.80-2.86]*** |
| Invasive ventilation | 120[5.6] | 49[1.9] | 3.06[2.18-4.28]*** |
| **In-hospital outcomes** |  |  |  |
| Recovered | 1813[85.3] | 2404[94.2] | 0.35[0.29-0.44]*** |
| Referred | 54[2.5] | 65[2.6] | 0.99[0.69-1.44] |
| Deaths | 258[12.1] | 82[3.2] | 4.16[3.22-5.37]*** |
| Odds ratio (OR) for categorical variables; Mean difference for continuous variables; CI confidence intervals; AST aspartate aminotransferase; ***p<0.001; **p<0.01; *p<0.05; |  |  |  |

**Table 2:** Clinical characteristics and outcomes according and to sex.

|  | Men (n=3386) | Women (n=1259) | Odds Ratio & Mean Difference<br>(95% CI) in men/women |
| --- | --- | --- | --- |
| Age: mean±SD | 45.5±17.8 | 47.1±18.5 | 1.60[0.43-2.76] |
| Low educational status | 800[23.4] | 624[49.8] | 0.31[0.27-0.36]*** |
| Household: mean±SD | 4.91±2.85 | 4.79±2.72 | -0.12[-0.03-0.06] |
| **Risk factors** |  |  |  |
| Smoking/tobacco | 1086[31.9] | 283[22.3] | 1.64[1.41-1.89]*** |
| Hypertension | 658[19.3] | 173[13.6] | 1.51[1.26-1.81]*** |
| Type 2 Diabetes | 666[19.6] | 111[8.7] | 2.53[2.05-3.13]*** |
| Thyroid disease | 27[0.8] | 11[0.9] | 0.91[0.45-1.84] |
| Coronary heart disease | 51[1.5] | 24[1.9] | 0.79[0.48-1.28] |
| Pulmonary disease | 135[4.0] | 58[4.6] | 0.85[0.63-1.18] |
| **Clinical findings** |  |  |  |
| Systolic BP mmHg | 125.1±11.9 | 126.0±12.9 | 0.88[0.09-1.66]* |
| SpO2 <90% | 252[11.4] | 105[12.2] | 0.88[0.69-1.12] |
| SpO2 90-94% | 397[18.0] | 164[19.1] | 0.89[0.73-1.08] |
| **Investigations** |  |  |  |
| Haemoglobin, g/dl | 12.7±2.3 | 12.6±2.2 | -0.10[-0.25-0.05] |
| White cells, 10<sup>9</sup> cells/L | 7.58±3.89 | 7.37±3.65 | -0.21[-0.46-0.04] |
| Lymphocyte:neutrophil | 0.36±0.27 | 0.35±0.46 | -0.01[-0.03-0.01] |
| Serum AST, units | 45.0±108.9 | 44.8±44.5 | -0.20[-6.40-6.00] |
| Sodium, mEq/L | 136.6±9.4 | 134.8±17.8 | -1.80[-2.59--1.01]*** |
| Creatinine, mg/dl | 0.94±0.47 | 0.97±0.56 | 0.03[-0.00-0.06] |
| **Clinical Outcomes** |  |  |  |
| Oxygen requirement | 600[17.6] | 261[20.6] | 0.77[0.65-0.90]** |
| Non-invasive ventilation | 236[6.9] | 98[7.7] | 0.88[0.69-1.33] |
| Invasive ventilation | 123[3.6] | 46[3.6] | 0.99[0.70-1.40] |
| **In-hospital outcomes** |  |  |  |
| Recovered | 3020[88.7] | 1197[94.3] | 0.95[0.39-0.65]*** |
| Referred | 104[3.0] | 15[1.2] | 0.77[0.54-1.09] |
| Deaths | 282[8.3] | 58[4.6] | 1.88[1.41-2.51]*** |
| Odds ratio (OR) for categorical variables; Mean difference for continuous variables; CI confidence intervals;<br>AST aspartate aminotransferase; ***p<0.001; **p<0.01; *p<0.05; |  |  |  |

Clinical characteristics, clinical findings, selected investigations and outcomes in patients with hypertension, diabetes and smoking/tobacco use are shown in Tables 3-5. Patients with known hypertension were older and had higher prevalence of diabetes, cardiovascular disease, hypothyroidism and smoking/tobacco (Table 3). Need for non-invasive ventilation was more (OR 1.82, CI 1.41-2.35) and deaths were significantly greater (OR 2.22, CI 1.74-2.83). Patients with diabetes were older, men with greater prevalence of tobacco, hypertension, admission BP, pulmonary disease and CV disease (Table 4). As compared to non-diabetics, oxygen requirement (OR 1.52, CI 1.26-1.82) and invasive ventilation (OR 1.64, CI 1.14-2.35) as well as deaths (OR 1.88, CI 1.46-2.43) were significantly greater. Patients with tobacco use (smokers, smokeless tobacco) had more men, with greater prevalence of hypertension, diabetes and CV disease (Table 5). Compared with non-tobacco users the need for oxygen (OR 1.23, CI 1.05-1.44,) non-invasive ventilation (OR 1.31, CI 1.04-1.66) and deaths (OR 1.29, CI 1.02-1.63) were significantly greater.

**Table 3:**
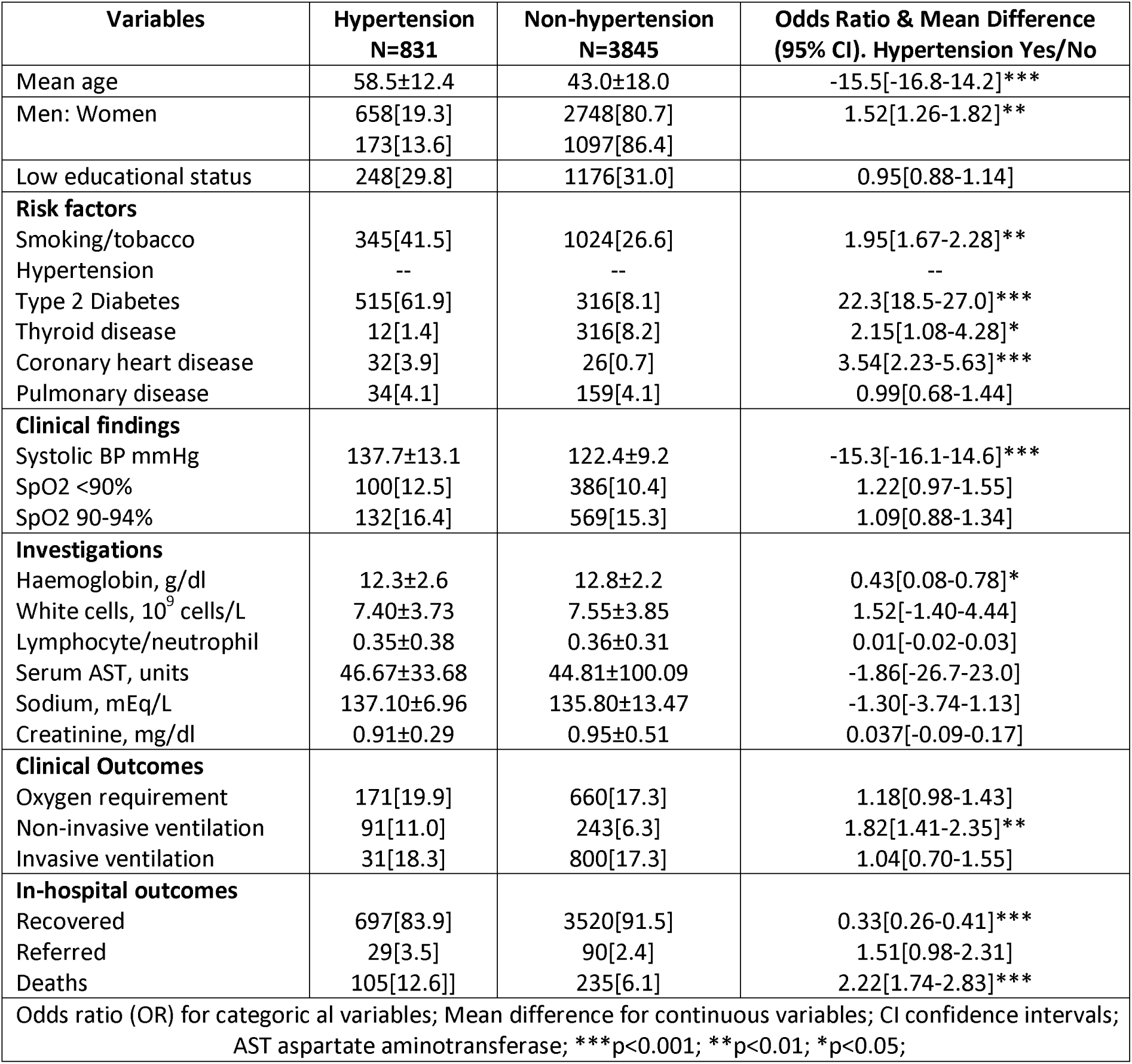
Clinical characteristics and outcomes according to hypertension.

**Table 4:** Clinical characteristics and outcomes according to diabetes.

| **Variables** | **Diabetes**<br>**N=777** | **Non-Diabetes**<br>**N=3899** | **Odds Ratio/ Mean Difference**<br>**(95% CI) Diabetes Yes/No** |
| --- | --- | --- | --- |
| Mean age | 57.6±12.9 | 43.4±18.1 | -14.2[-15.5-12.9]*** |
| Men: Women | 549[16.1]<br>228[18.0] | 2857[83.9]<br>1042[82.0] | 0.53[0.71-1.00]* |
| Low educational status | 220[28.3] | 1204[31.3] | 0.87[0.74-1.03] |
| **Risk factors** |  |  |  |
| Smoking/tobacco | 303[39.0] | 1066[27.4] | 1.70[1.45-1.99]** |
| Hypertension | 515[66.3] | 316[8.1] | 22.3[18.5-27.0]*** |
| Type 2 Diabetes | -- | -- | -- |
| Thyroid disease | 10[1.3] | 28[0.7] | 1.80[0.87-3.73] |
| Coronary heart disease | 27[3.5] | 48[1.2] | 2.89[1.79-4.66]*** |
| Pulmonary disease | 48[6.2] | 145[3.7] | 1.70[1.22-2.38]** |
| **Clinical findings** |  |  |  |
| Systolic BP mmHg | 135.1±13.7 | 123.1±10.0 | -11.9[-12.8-11.1]*** |
| SpO2 <90% | 92[11.84] | 394[10.10] | 1.19[0.94-1.52] |
| SpO2 90-94% | 119[15.31] | 582[14.92] | 1.03[0.83-1.28] |
| **Investigations** |  |  |  |
| Haemoglobin, g/dl | 12.3±2.4 | 12.8±2.6 | 0.48[0.13-0.83]** |
| White cells, 10<sup>9</sup> cells/L | 7.38±3.81 | 7.55±3.83 | 1.75[-1.26-4.78] |
| Lymphocyte/neutrophil | 0.35±0.39 | 0.36±0.31 | 0.01[-0.02-0.02] |
| Serum AST, units | 42.93±35.83 | 45.12±100.58 | 2.19[-21.04-25.45] |
| Sodium, mEq/L | 137.12±6.08 | 135.82±13.44 | -1.29[-3.83-1.24] |
| Creatinine, mg/dl | 0.93±0.32 | 0.95±0.51 | 0.02[-0.09 to 0.14] |
| **Clinical Outcomes** |  |  |  |
| Oxygen requirement | 187[24.1] | 674[17.3] | 1.52[1.26-1.82]*** |
| Non-invasive ventilation | 64[8.2] | 270[6.9] | 1.21[0.91-1.60] |
| Invasive ventilation | 41[5.3] | 128[3.3] | 1.64[1.14-2.35]** |
| **In-hospital outcomes** |  |  |  |
| Recovered | 653[84.0] | 3564[91.4] | 0.49[0.39-0.62]*** |
| Referred | 35[4.5] | 84[2.2] | 2.14[1.43-3.20]*** |
| Deaths | 89[11.5] | 251[6.4] | 1.88[1.46-2.43]*** |
| Odds ratio (OR) for categorical variables; Mean difference for continuous variables; CI confidence intervals;<br>AST aspartate aminotransferase; ***p<0.001; **p<0.01; *p<0.05; |  |  |  |

**Table 5:** Clinical characteristics and outcomes according to smoking/tobacco use.

| **Variables** | **Smoking/tobacco**<br>**N=1369** | **Non-smoker/tobacco**<br>**N=3307** | **Odds Ratio/ Mean Difference**<br>**(95% CI) Tobacco Yes/No** |
| --- | --- | --- | --- |
| Mean age | 46.3±18.1 | 45.5±18.1 | -0.86[-2.00-0.28] |
| Men: Women | 1086[79.3]<br>283[20.7] | 2320[70.2]<br>987[29.8] | 1.64[1.41-1.89]*** |
| Low educational status | 414[30.6] | 1010[30.8] | 1.01[0.88-1.16] |
| **Risk factors** |  |  |  |
| Smoking/tobacco | -- | -- | -- |
| Hypertension | 345[25.2] | 486[14.7] | 1.96[1.67-2.28]*** |
| Type 2 Diabetes | 303[22.1] | 474[14.3] | 1.69[1.45-1.99]*** |
| Thyroid disease | 5[0.4] | 33[1.0] | 0.36[0.14-0.93]* |
| Coronary heart disease | 34[2.5] | 41[1.2] | 2.03[1.28-3.21]** |
| Pulmonary disease | 67[4.9] | 126[3.8] | 1.29[0.96-1.76] |
| **Clinical findings** |  |  |  |
| Systolic BP mmHg | 126.1±12.6 | 124.7±11.2] | -1.3[-2.1-0.6]*** |
| SpO2 <90% | 131[9.9] | 355[11.1] | 0.88[0.71-1.08] |
| SpO2 90-94% | 194[14.7] | 507[15.8] | 0.91[0.76-1.09] |
| **Investigations** |  |  |  |
| Haemoglobin, g/dl | 12.7±2.1 | 12.7±2.3 | -0.03[-0.28-0.22] |
| White cells, 10<sup>9</sup> cells/L | 7.49±3.73 | 7.54±3.87 | 0.05[-0.19-3.02] |
| Lymphocyte/neutrophil | 0.34±0.25 | 0.36±0.35 | 0.02[0.00-0.04]* |
| Serum AST, units | 56.78±85.45 | 40.74±46.51 | -16.0[-30.6-1.5] |
| Sodium, mEq/L | 136.15±14.26 | 136.01±11.49 | -0.15[-2.22-1.92] |
| Creatinine, mg/dl | 0.95±0.46 | 0.95±0.51 | -0.01[-0.82-0.07] |
| **Clinical Outcomes** |  |  |  |
| Oxygen requirement | 283[20.7] | 578[17.5] | 1.23[1.05-1.44]* |
| Non-invasive ventilation | 116[8.5] | 218[6.6] | 1.31[1.04-1.66]* |
| Invasive ventilation | 46[3.4] | 123[3.7] | 0.90[0.64-1.27] |
| **In-hospital outcomes** |  |  |  |
| Recovered | 1223[89.3] | 2994[90.5] | 0.87[0.71-1.08] |
| Referred | 29[2.2] | 90[2.7] | 0.77[0.51-1.18] |
| Deaths | 117[8.5] | 223[6.7] | 1.29[1.02-1.63]* |
| Odds ratio (OR) for categorical variables; Mean difference for continuous variables; CI confidence intervals;<br>AST aspartate aminotransferase; ***p<0.001; **p<0.01; *p<0.05; |  |  |  |

We performed age- and sex-adjusted and multivariate analyses to determine association of various CV risk factors with COVID-19 related outcomes (Table 6). Age ≥50 years emerged as the most important risk factor with significantly greater deaths on univariate (OR 4.16, 95% CI 3.22-5.37)), sex-adjusted (OR 4.15, 95% CI 3.21-5.35) as well as multivariate analyses (OR 3.89, 95% CI 2.97-5.10). On univariate analyses, male sex (OR 1.88, 1.41-2.51), hypertension (2.22, 1.74-2.83), diabetes (1.88, 1.46-2.43) and tobacco (1.29, 1.02-1.63) were associated with more deaths (Figure 2, p<0.001). There was moderate attenuation of significance with age and sex adjusted analyses, but hypertension (1.32, 1.02-1.70) and tobacco use (1.28, 1.00-1.63) continued to be significant. Following multivariate analyses significance of all the risk factors completely attenuated (Figure 2). In patients age ≥50 years compared with the younger, the need for invasive ventilation as well as non-invasive ventilation were higher (Table 5). Hypertension was significantly associated with greater risk of invasive ventilation on univariate and adjusted analyses and greater risk of non-invasive ventilation on univariate analyses. Diabetes patients had greater risk of non-invasive and invasive ventilation on univariate analyses which attenuated on age-sex adjusted and multivariate analyses (Table 6).

**Table 6:** Univariate and multivariate logistic regression analyses (odds ratio and 95% confidence intervals) for adverse outcomes in various cardiovascular risk factor groups.

|  | **Risk factor variables** | **Unadjusted Odds Ratios** | **Age and Sex adjusted** | **Plus household size, risk factors, comorbidities** |
| --- | --- | --- | --- | --- |
| Deaths | Age >=50y | 4.16[3.22-5.37] | 4.15[3.21-5.35]<sup>§</sup> | 3.89[2.97-5.10] |
|  | Men: Women | 1.88[1.41-2.51] | 1.26[0.91-1.48]<sup>&</sup> | 1.05[0.82-1.35] |
|  | Hypertension | 2.22[1.74-2.83] | 1.32[1.02-1.70] | 1.27[0.94-1.73] |
|  | Diabetes | 1.88[1.46-2.43] | 1.16[0.89-1.52] | 1.02[0.75-1.40] |
|  | Tobacco | 1.29[1.02-1.63] | 1.28[1.00-1.63] | 1.23[0.96-1.57] |
| Invasive ventilation | Age >=50y | 3.06[2.18-4.28] | 3.06[2.18-4.29] | 1.29[0.73-2.29] |
|  | Men: Women | 1.00[0.71-1.42] | 0.94[0.67-1.33] | 1.09[0.76-1.55] |
|  | Hypertension | 3.05[2.22-4.20] | 2.12[1.51-2.97] | 2.64[1.78-3.92] |
|  | Diabetes | 1.64[1.14-2.35] | 1.12[0.77-1.62] | 0.64[0.42-0.99] |
|  | Tobacco | 1.27[0.92-1.76] | 1.24[0.89-1.72] | 1.12[0.79-1.57] |
| Non-invasive ventilation | Age >=50y | 2.27[1.80-2.86] | 2.26[1.79-2.85] | 2.00[1.57-2.55] |
|  | Men: Women | 0.88[0.69-1.33] | 0.93[0.73-1.19]<sup>l</sup> | 0.92[0.65-1.31] |
|  | Hypertension | 1.82[1.41-2.35] | 1.29[0.99-1.69] | 0.91[0.63-1.32] |
|  | Diabetes | 1.21[0.91-1.60] | 0.86[0.64-1.15] | 0.49[0.34-0.69] |
|  | Tobacco | 1.31[1.04-1.66] | 1.32[1.04-1.67] | 0.89[0.69-1.14] |

**Figure 2:**
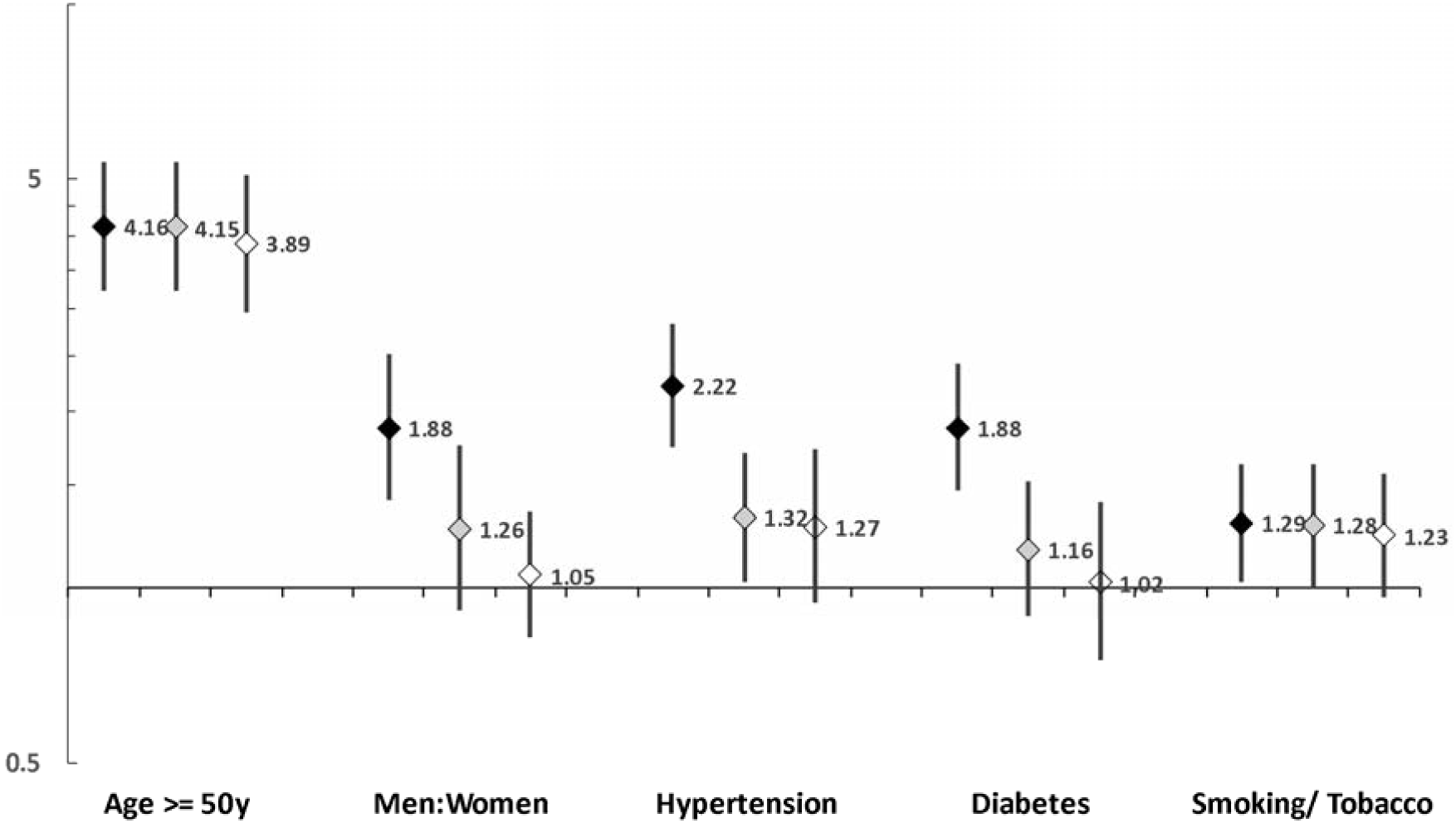
Odds ratio and 95% confidence intervals for COVID-19 related deaths in patients in old vs young, men vs women, hypertension, diabetes and tobacco groups on univariate (black markers), age and sex adjusted (grey markers) and multivariate (open markers) logistic regression

## DISCUSSION

This study shows that modifiable and non-modifiable cardiovascular risk factors-hypertension, diabetes, tobacco use, increasing age and male sex are associated with greater risk of death and adverse outcomes among hospitalized COVID-19 patients. Significance of hypertension and tobacco is maintained even after age- and sex-adjustment highlighting importance of these factors.

Our results are similar to most of the previous meta-analyses that have identified age as the most important risk factor for adverse outcomes in COVID-19.^2-5^ Cardiovascular risk factors-hypertension and diabetes-have been identified as important in most previous studies.^3-5^ In the present study, although both of these factors are associated with greater risk of death and some adverse outcomes (Table 6), there is substantial attenuation after age-adjustment for hypertension and complete attenuation in diabetes indicating age and sex as intermediate pathways of increased risk. A study limitation is that we included COVID-19 patients with known hypertension and diabetes at time of admission as risk factor. Given the fact that in India only about half of the patients with hypertension and two-thirds with diabetes are aware of their condition,^19^ the prevalence of these conditions might have been higher in our cohort. However, many individuals with hypertension present with low BP in acute COVID-19 and therefore estimation of prevalence of hypertension based on measured BP would have been erroneous. Moreover, our unadjusted OR of 2.22 (CI 1.74-2.51) and age-sex adjusted OR of 1.32 (CI 1.02-1.70) is similar to many previous studies and meta-analyses have calculated hypertension related OR in COVID-19 between 1.90 (CI 1.69-2.35)^3^ and 2.50 (CI 2.15-2.90),^5^ similar to the present study. We did not inquire the type of anti-hypertensive patients in our study cohort. Certain BP medications such as renin-angiotensin system blockers are known to be useful in COVID-19.^20,21^

Previous meta-analyses including studies from India have identified diabetes as equally important as hypertension for adverse COVID-19 related outcomes.^11,12^ In the present study the unadjusted OR for diabetes and deaths were 1.88 (CI 1.46-2.43), however, the risk significantly attenuated after age and sex adjustment to OR 1.16 (CI 0.89-1.52) which is different from the previous studies. In the present study, we included patients with known diabetes only and this is a study limitation.^22^ It is likely that using biomarkers for diabetes diagnosis (HbA1c, glucose tolerance test, etc.) we would have diagnosed more diabetics, but these criteria are fraught with inconsistency during any acute illness. We also found significant association of smoking/tobacco use with death and other adverse outcomes in our cohort. This association holds even after multivariate adjustments and shows that tobacco is an important risk factor. This is different from many previous studies that have reported disparate results.^23^ On the other hand, a large meta-analysis that included 109 articles and 517,020 patients reported that smoking was associated with increased risk of admission to ICU and increased mortality (OR 1.58, CI 1.38-1.81).^24^ Meta-regression analyses identified that the increased risk of smoking was mediated via increased age, hypertension and diabetes. Our finding is similar to data from Chinese cohorts (high rates of smoking) in the aforementioned meta-analysis.^24^

The study has several other limitations. This is a single-centre study and the results may not be externally valid within India or other countries as Rajasthan is one of the less-developed states in the nation with lower prevalence of hypertension and diabetes.^19,25^ On the other hand, this is the largest study from India and much larger than many other studies from developed countries, the data were obtained from a government hospital thus assuring wider population representation and better data granularity. Secondly, this is not a population-based study as many studies from Europe and North America are,^7^ and we may have missed data on milder forms of disease. Thirdly, we do not have data on obesity or body-mass index which is an important COVID-19 risk factor in hospital- and population-based studies.^7,24^ Fourthly, we also do not have data on biochemical investigations for all the patients, although data on white cell count are available for more than 90%. Also, we do not have data of radiological evaluation of all the patients as it is well known that computerized tomographic images provide important prognostic information.^26^ Fifthly, the rate of progression of illness as well as greater details of causes of deaths are not available and this is a study limitation as discussed earlier.^17^ We did not analyse data on patients with known cardiovascular disease, cancers and chronic kidney disease because of small numbers of these patients (Figure 1). And finally, we did not obtain data regarding Post-COVID syndrome which is emerging as important health problem especially in persons with comorbidities.^27^ On the other hand, this is the largest study from India and with robust data has important clinical implications.

In conclusion, this study shows that older patients, males, and those with hypertension, diabetes and any tobacco use have greater risk of deaths and adverse outcomes from COVID-19. These individuals should be educated regarding the increased risk and should aggressively follow all non-pharmacological physical measures for prevention.^28^ These groups should also be prioritized for vaccinations.^29^ Clinicians are advised to seek early evidence of deterioration of pulmonary function and signs of cardiovascular and extrapulmonary manifestation of acute COVID-19 in these patients and provide optimum management.^30,31^ It is likely that with proper preventive and therapeutic interventions higher risk of the patients with CV risk factors in COVID-19 can be mitigated.

## Data Availability

All the available data are included in the article.

## Notes

### Competing Interest Statement

The authors have declared no competing interest.

### Clinical Trial

ctri.nic.in REF/2020/06/034036

### Funding Statement

No funding has been received for this work.

### Author Declarations

Rajasthan University of Health Sciences, Jaipur, India. Institutional Ethics Committee: CDSCO Registration Number: CR/762/Inst/RJ/2015.

## REFERENCES

1. Nishiga M, Wang DW, Lewis DB, Wu JC. COVID-19 and cardiovascular disease: from basic mechanisms to clinical perspectives. Nature Rev Cardiol. 2020; 543–558.

2. Matsushita K, Ding N, Kou M, et al. The relationship of COVID-19 severity with cardiovascular disease and its traditional risk factors: a systematic review and meta-analysis. Global Heart. 2020; 15:e64.

3. Chidambaram V, Tun NL, Haque WZ, et al. Factors associated with disease severity and mortality among patients with COVID-19: a systematic review and meta-analysis. PLoS One. 2020; 15:e0241541.

4. Silverio A, Maio MD, Citro R, et al. Cardiovascular risk factors and mortality in hospitalized patients with COVID-19: a systematic review and meta-analysis of 45 studies and 18300 patients. BMC Cardiovasc Disord. 2021; 21:e23.

5. Bae SA, Kim SR, Kim MN, Shim WJ, Park SM. Impact of cardiovascular disease and risk factors on fatal outcomes in patients with COVID-19 according to age: a systematic review and meta-analysis. Heart. 2021; 107:373–380.

6. Collard D, Nurmohamed NS, Kaiser Y, et al. Cardiovascular risk factors and COVID-19 outcomes in hospitalized patients: a prospective cohort study. BMJ Open. 2021; 11:e045482.

7. Gao M, Piernas C, Astbury NM, et al. Associations between body mass index and COVID-19 severity in 6.9 million people in England: a prospective, community-based, cohort study. Lancet Diabetes Endocrinol. 2021; 9:350–359.

8. Ritchie H, Ortiz-Ospina E, Beltekian D, et al. Coronavirus pandemic (COVID-19). Available at: https://ourworldindata.org/coronavirus. Accessed May 19, 2021.

9. Global Burden of Diseases CVD Collaborators. Global burden of cardiovascular diseases and risk factors, 1990-2019: Update from the Global Burden of Disease 2019 Study. J Am Coll Cardiol. 2020; 76:2982–3021.

10. Ritchie H, Ortiz-Ospina E, Beltekian D, et al. India: Coronavirus Pandemic Country Profile. Available at: https://ourworldindata.org/coronavirus/country/india. Accessed May 5, 2021.

11. Singh AK, Gilles CL, Singh R, et al. Prevalence of co-morbidities and their association with mortality in patients with COVID-19: a systematic review and meta-analysis. Diabetes Obes Metab. 2020; 22:1915–1924.

12. Nandy K, Salunke A, Pathak SK, et al. Coronavirus disease (COVID-19): a systematic review and meta-analysis to evaluate the impact of various comorbidities and serious events. Diabetes Metab Syndr. 2020; 14:1017–1025.

13. Chakafana G, Mutithu D, Hoevelmann J, Ntusi N, Sliwa K. Interplay of COVID-10 and cardiovascular diseases in Africa: an observational snapshot. Clin Res Cardiol. 2020; 109:1460–1468.

14. Gaur K, Khedar RS, Mangal K, Sharma AK, Dhamija RK, Gupta R. Macrolevel association of COVID-19 with non-communicable disease risk factors in India. Diabetes Metab Syndr. 2021; 15:343–350.

15. Sharma AK, Ahmed A, Baig VN, et al. Characteristics and outcomes of hospitalized young adults with mild to moderate COVID-19 at a university hospital in India. J Assoc Physicians India. 2020; 68(8):62–65.

16. Sharma S, Sharma AK, Dalela G, et al. Association of SARS CoV-2 cycle threshold (Ct) with clinical outcomes: a hospital-based study. J Assoc Physicians India. 2021; 69(6):86–90.

17. Sharma AK, Gupta R, Baig VN, et al. Socioeconomic status and COVID-19 related outcomes in India: A hospital-based study. medRxiv preprints. 2021; DOI 10.1101/2021.05.17.21257364.

18. Government of India, Ministry of Health and Family Welfare. Clinical management protocol: COVID-19. Available at: http://www.rajswasthya.nic.in/PDF/COVID%20-19/FOR%20HOSPITALS/27.06.2020.pdf. Accessed 30 April 2021.

19. Gupta R, Gaur K, Ram CVS. Emerging trends in hypertension epidemiology in India. J Human Hypertens. 2019. 33:575–587.

20. Savoia C, Volpe M, Kreutz R. Hypertension, a moving target in COVID-19: current views and perspectives. Circ Res. 2021; 128:1062–1079.

21. Tavares CAM, Bailey MA, Girardi ACC. Biological context linking hypertension and higher risk for COVID-19 severity. Front Physiol. 2020; 11:599729.

22. Gupta A, Gupta R, Sharma KK, et al. Prevalence of diabetes and cardiovascular risk factors in middle-class urban populations in India. BMJ Open Diabetes Res Care. 2014; 2:e000048.

23. Dorjee K, Kim H, Bonomo E, Dolma R. Prevalence and predictors of death and severe disease in patients hospitalized due to COVID-19: a comprehensive systematic review and meta-analysis of 77 studies and 38,000 patients. PLoS One. 2020; 15:e0243191.

24. Zhang H, Ma S, Han T, et al. Association of smoking history with severe and critical outcomes in COVID-19 patients: a systematic review and meta-analysis. Eur J Intergr Med. 2021; 43:101313.

25. Gupta R, Gaur K. Epidemiology of ischemic heart disease and diabetes in India: An overview of the twin epidemic. Current Diabetes Reviews. 2020; 17: EPub.

26. Machnicki S, Patel D, Singh A, et al. The usefulness of chest CT imaging in patients with suspected or diagnosed COVID-19: a review of literature. Chest. 2021; EPub.

27. Nalbandian A, Sehgal K, Gupta A, et al. Post-acute COVID-19 syndrome. Nature Med. 2021; 27:601–615.

28. Kucharski A, Klepac P, Conlan AJ, et al. Effectiveness of isolation, testing, contact tracing, and physical distancing on reducing transmission of SARS-CoV-2 in different settings: a mathematical modelling study. Lancet Infect Dis. 2020; 20:1151–1160.

29. Schmidt H, Weintraub R, Williams MA, et al. Equitable allocation of COVID-19 vaccines in the United States. Nature Med. 2021; EPub.

30. Azevodo RB, Botelho BG, de Hollanda JVG, et al. COVID-19 and the cardiovascular system: a comprehensive review. J Human Hypertens. 2021; 35:4–11.

31. Gupta A, Madhavan MV, Sehgal K, et al. Extrapulmonary manifestations of COVID-19. Nature Med. 2020; 26:1017–1032.

